# Automating neoantigen selection for personalized cancer vaccine design

**DOI:** 10.64898/2026.06.24.26356293

**Authors:** Jennie X. Yao, Kartik Singhal, Susanna Kiwala, Evelyn Schmidt, S. Peter Goedegebuure, Christopher A. Miller, Huiming Xia, Kelsy C. Cotto, Adam Coffman, My H. Hoang, Mariam Khanfar, Jinglun Li, Luke Hendrickson, Isabel Risch, Sherri R. Davies, Feiyu Du, Gue Su Chang, Jasreet Hundal, Jeffrey P. Ward, William B. Inabinett, William A. Hoos, Tanner M. Johanns, Gavin P. Dunn, Russell K. Pachynski, Todd A. Fehniger, Jennifer A. Foltz, William E. Gillanders, Malachi Griffith, Obi L. Griffith

**Affiliations:** Division of Oncology, Department of Medicine, Washington University School of Medicine, St. Louis, MO; Siteman Cancer Center, Washington University School of Medicine, St. Louis, MO; McDonnell Genome Institute, Washington University School of Medicine, St. Louis, MO; Department of Surgery, Washington University School of Medicine, St. Louis, MO; Department of Genetics, Washington University School of Medicine, St. Louis, MO, USA; Jaime Leandro Foundation for Therapeutic Cancer Vaccines, Chapel Hill, NC, USA; Department of Pathology and Immunology, Washington University School of Medicine, St Louis, MO, USA; Department of Neurosurgery, Massachusetts General Hospital/Mass General Brigham, Boston, MA

## Abstract

Advancements in immunogenomics and immuno-oncology have enabled the development of personalized cancer vaccines (PCVs) that target cancer cell-specific somatic variants. A subset of these variants produce neoantigens that, when presented on tumor cells by MHC molecules, have the potential to elicit a robust and specific immune response. To date, there are over one hundred interventional studies listed on *clinicaltrials.gov* that explore the use of PCVs. We have supported a number of these trials through the creation of bioinformatic pipelines, tools, and procedures for the identification of patient-specific neoantigen candidates. While many of these steps have been automated, the final selection of neoantigen candidates often relies on expert manual review, creating a bottleneck that limits scalability and full automation of PCV workflows. Addressing this challenge, we introduce NEAT (Neoantigen Evaluation & Automated Triage), a machine learning-based approach that enables automated neoantigen candidate prioritization and supports the transition toward more scalable and reproducible PCV design. We implemented a prediction model trained and tested on existing vaccine design results from 33 patients and 1,943 peptides, across 3 clinical trials, including 439 peptides prioritized for PCV inclusion. This model uses features such as tumor variant allele frequency, RNA expression, driver gene status, binding/presentation scores, and transcript support level to automatically predict whether a peptide will be accepted, rejected, or require further human review before inclusion in a vaccine. The model achieved a sensitivity of 0.847 and specificity of 0.924, with an area under the curve of 0.955. The model predictions have been incorporated in pVACtools v7.0.0. By integrating this model into the vaccine development pipeline, we foresee a significant reduction in the time required to transition from patient sample collection to vaccine manufacturing, thereby enhancing the efficiency and scalability of PCV production.

## Introduction

Personalized cancer vaccines (PCVs) are a promising approach to cancer immunotherapy that leverage cancer-specific genetic mutations found in tumor cells to stimulate an immune response^1,2^. These vaccines are designed to target neoantigens, which are peptides derived from a subset of somatic mutations and are absent from normal tissues, thus minimizing the risk of attacking healthy cells. Neoantigens that are successfully processed and presented on the cell surface by class I and class II major histocompatibility complexes (MHC; human leukocyte antigens, HLA) have the potential to be recognized by CD8+ and CD4+ T cells and elicit anti-tumor immune responses^3,4^. Recent studies have shown promising results regarding the safety and efficacy of PCVs in several cancer types, including melanoma^5,6^, glioblastoma^7,8^, hepatocellular carcinoma^9^, triple-negative breast cancer^10^, and follicular lymphoma^11^.

The development of treatment options to target neoantigens requires the identification of neoantigens that are recognizable by T cells. However, only a small subset of candidate neoantigens are presented at the cell’s surface and capable of inducing a potent immunogenic response, making their identification challenging and a priority for tool development. The immunogenicity of a neoantigen depends on a complex series of biological events, including variant expression, proteasomal processing, transport and presentation at the cell’s surface, and the ability to elicit a T-cell response. To this end, various computational tools, leveraging advances in next generation sequencing (NGS) and bioinformatics, have been developed to predict immunogenic neoantigens. Using these tools, many steps of the neoantigen selection process have been automated.

Despite these advances, final decision-making often remains a manual and expert-driven process often carried out by an Immunogenomics Tumor Board (ITB) or similar multidisciplinary team including physician-scientists, and specialists in immunology, genomics, and bioinformatics (Figure 1). During ITB review, members evaluate each candidate based on a variety of features, including predicted MHC binding affinity across multiple algorithms and the level of agreement among them; whether the mutated peptide aligns with or diverges from the reference proteome; and whether the mutation occurs in a known cancer driver gene. Candidates are also subject to manual inspection, for example in the Integrative Genomics Viewer (IGV), to assess transcript structure, splicing patterns, and the presence of nearby variants that may affect interpretation, and to verify read coverage, particularly for complex variants. While this expert-driven process enables comprehensive evaluation, it also introduces challenges, including potential inconsistency in the application of selection criteria across patients and meetings, difficulty in validating decisions derived from subjective judgment rather than standardized criteria, and increased risk of human error when integrating complex, multifactorial evidence. These challenges are particularly relevant as PCV advances toward broader clinical implementation, where reproducibility, traceability, and standardized decision-making are increasingly important for regulatory review and quality assurance. As efforts to evaluate PCVs expand and more patients are enrolled into clinical trials, each with potentially hundreds of candidate neoantigens, the number of neoantigens requiring ITB and manual review has grown substantially. This increase in review volume has led to longer turnaround times, creating a bottleneck that limits the scalability of PCV development.

**Figure 1.**
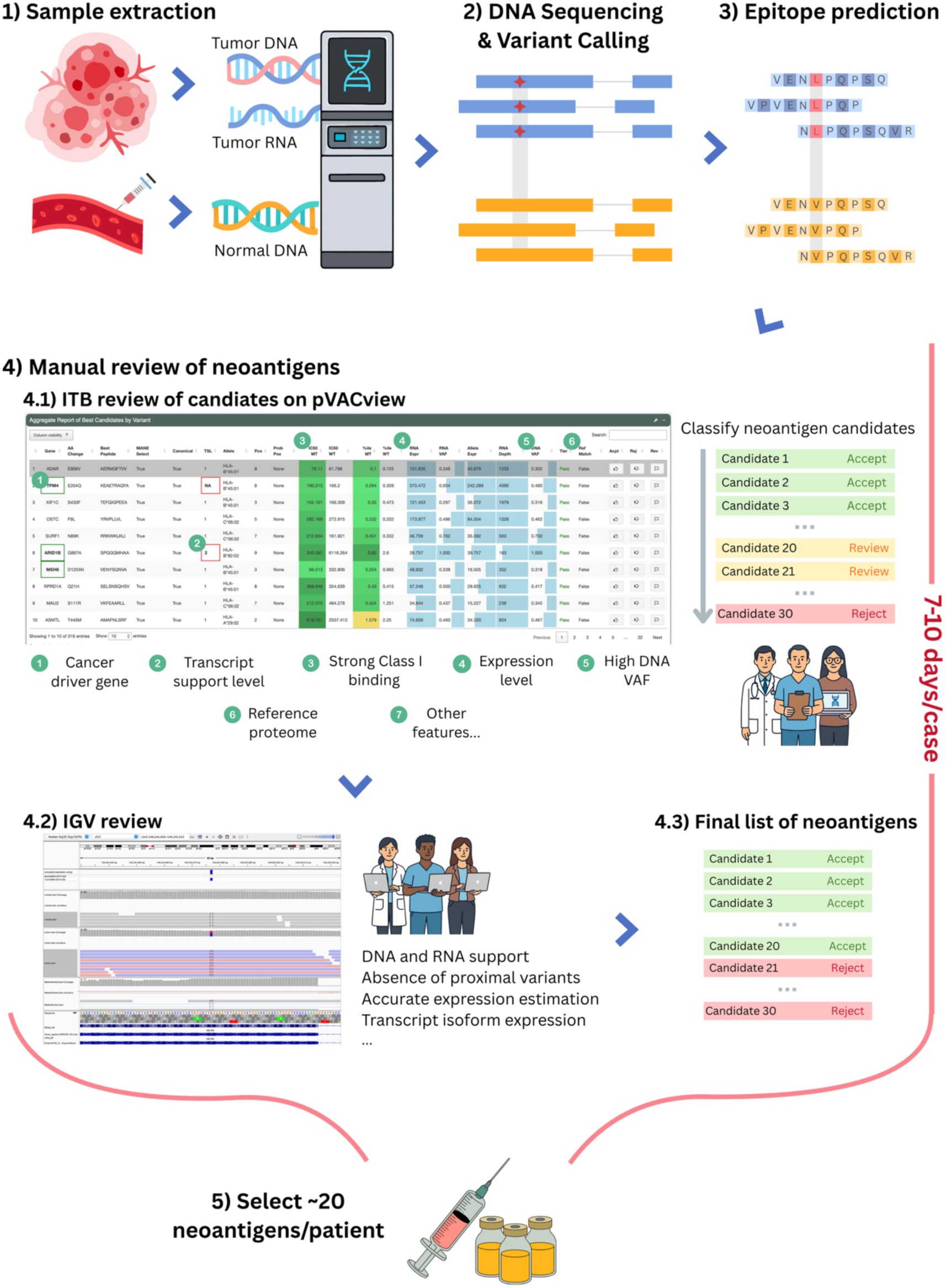
Neoantigen prioritization workflow overview. Overview of the PCV design process. 1) Tumor DNA, tumor RNA, and matched normal DNA are extracted from patient samples. 2) DNA sequencing and variant calling are performed to identify somatic mutations through comparison of tumor and normal sequences. 3) Candidate neoantigen epitopes are predicted computationally, generating a set of peptide sequences for downstream evaluation. 4.1) Members of the ITB perform manual review of each candidate using the pVACview interface, considering features such as cancer driver gene status, transcript support level, MHC Class I binding affinity, expression level, tumor DNA variant allele frequency (VAF), and similarity to the human reference proteome, among other criteria. Candidates are assigned an evaluation status (Accept, Review, or Reject) and prioritized accordingly. 4.2) Selected candidates undergo further manual inspection using the IGV to examine supporting evidence from DNA and RNA. 4.3) The ITB review and IGV inspection together inform a final curated list of neoantigen candidates. This expert review process typically takes 7–10 days per case. 5) Approximately 20 neoantigens per patient are ultimately selected for PCV manufacturing.

Cancer research is increasingly driven by the integration of large-scale, multidimensional datasets. Artificial intelligence (AI), particularly machine learning (ML), plays a central role in this transformation by enabling data-driven decision support systems for precision medicine. With the growing availability of datasets containing thousands of neoantigen selection outcomes, ML methods offer a powerful means to capture the multidimensional structure of neoantigen features and learn predictive models for candidate prioritization. In this study, we developed and integrated NEAT (Neoantigen Evaluation & Automated Triage), a random forest–based ML model, into our previously established pVACtools suite^12–15^ (Figure 2). The model demonstrates performance comparable to manual expert review while significantly improving consistency, efficiency, and scalability across diverse cancer types. In addition to automating the selection process, the model provides interpretable insights into key features that drive neoantigen immunogenicity, enhancing our understanding of optimal vaccine design.

**Figure 2.**
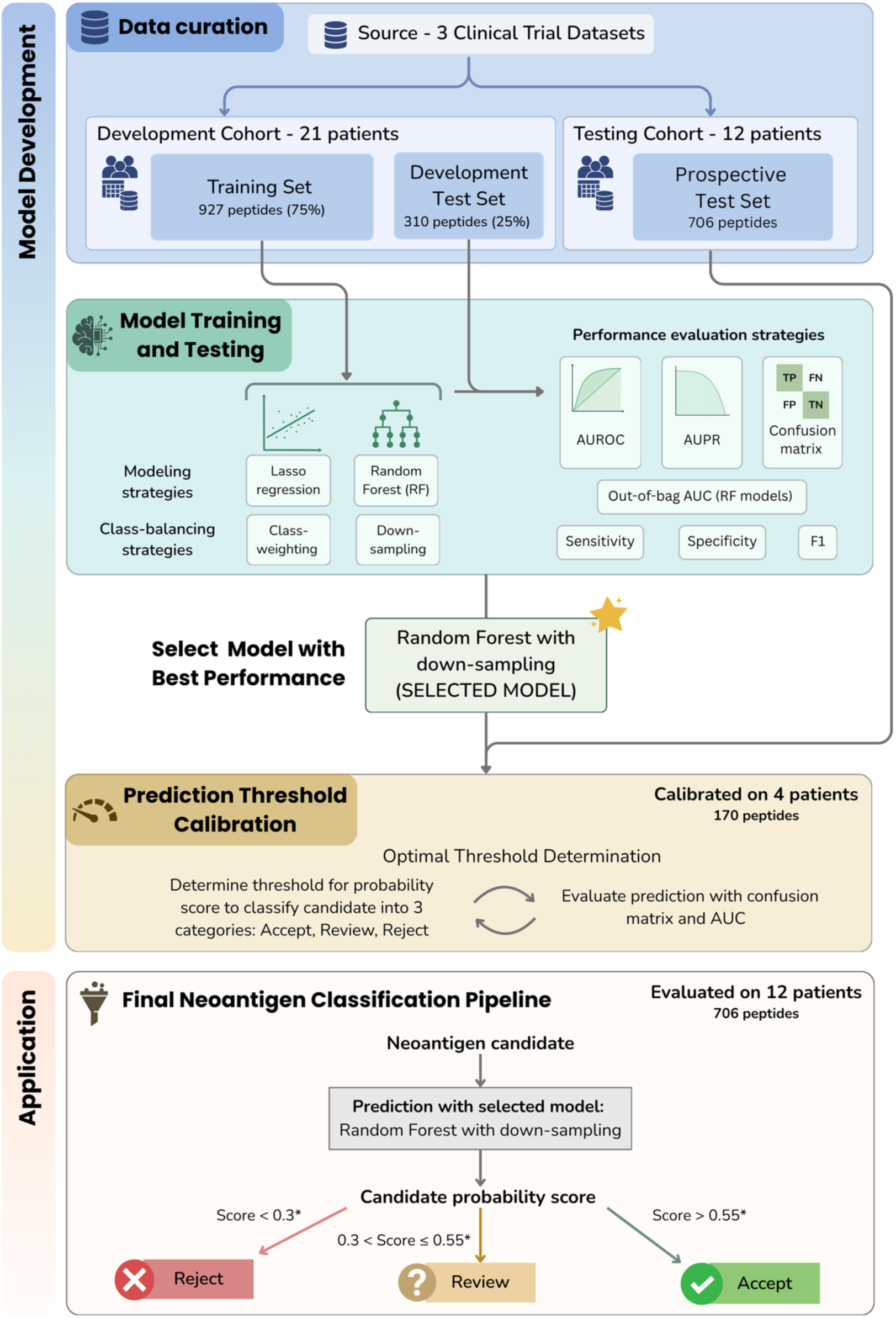
Machine learning model development workflow and clinical application. The workflow is divided into two phases: Model Development, in which candidate models were trained, evaluated, and a final model selected (Random forest with down-sampling), followed by probability threshold calibration on 4 patients (170 peptides); and Application, in which the selected model was applied to the full prospective test set (12 patients, 706 peptides) to classify each neoantigen candidate as Accept, Review, or Reject. *Thresholds (0.3 and 0.55) were determined during the calibration step and are fully adjustable to suit different clinical contexts.

## Methods

### Data

To build a prediction model to effectively prioritize neoantigen candidates, we compiled a list of neopeptides that were carefully prioritized and labeled (accept, reject, or review) in past ITB meetings. These neopeptides were generated for cancer patients selected from three ongoing or completed clinical trials at Washington University: a pancreatic cancer cohort (NCT05111353), a metastatic triple-negative breast cancer (TNBC) cohort (NCT03606967)^10^, and a basket trial that contains patients with multiple types of cancer (NCT05741242).

The final training and development test dataset included 1,237 peptides, of which 927 (75%) were used for training and 310 (25%) were used for testing (Table 1.1). Of all of the peptide candidates, 297 were accepted by our ITB to be included in a patient’s final vaccine and 940 were rejected. An additional prospective test set of 706 peptides from 12 patients was included for threshold calibration and prediction performance evaluation. Of these, 142 were accepted by ITB, 476 were rejected, and 88 were marked for further review (Table 1.2).

**Table 1.1.** Trial and tumor type breakdown of training set and development test set peptides.

| Trial | Tumor type | # of Patients | # of peptides in training set |  | # of peptides in development test set |  |
| --- | --- | --- | --- | --- | --- | --- |
|  |  |  | Accept | Reject | Accept | Reject |
| NCT05111353 | Pancreatic cancer | 7 | 68 | 153 | 28 | 45 |
| NCT03606967 | Metastatic TNBC | 6 | 74 | 294 | 29 | 84 |
| NCT05741242 | Osteosarcoma | 1 | 9 | 16 | 3 | 4 |
|  | Lung adenocarcinoma | 2 | 23 | 115 | 5 | 47 |
|  | Non-Small Cell Lung Adenocarcinoma | 1 | 7 | 59 | 9 | 18 |
|  | Synovial Sarcoma | 1 | 7 | 13 | 2 | 1 |
|  | Pancreatic cancer | 2 | 15 | 49 | 7 | 16 |
|  | Glioblastoma | 1 | 9 | 16 | 2 | 10 |
|  | Total | 21 | 212 | 715 | 85 | 225 |
|  |  |  | 927 |  | 310 |  |
|  |  |  | 1237 |  |  |  |

**Table 1.2.** Trial and tumor type breakdown of prospective test set peptides.

| Trial | Tumor type | # of Patients | # of Peptides |  |  |
| --- | --- | --- | --- | --- | --- |
|  |  |  | Accept | Reject | Review |
| NCT05111353 | Pancreatic cancer | 7 | 68 | 170 | 47 |
| NCT03606967 | Metastatic TNBC | 5 | 74 | 306 | 41 |
|  | Total | 12 | 142 | 476 | 88 |
|  |  |  | 706 |  |  |

### Features

The genomic data of all patients was preprocessed with the pVACtools^13^ pipeline as part of the ImmunoNX protocol^15^, which integrates data from a patient’s normal whole exome sequence (WES), tumor WES, tumor RNA-seq, and HLA haplotype to generate a list of candidate peptides that may be presented by tumor cells. For our ML model, we included a total of 72 features (Supplementary Table 1). We included features that are relevant to mutation qualities such as the tumor DNA and RNA variant allele frequencies (VAF), whether the mutated gene is a driver for the cancer type, transcript support level (TSL), and whether the peptide is found in the human reference proteome. Binding affinity and presentation of a peptide to MHC class I or class II are key features when prioritizing, and for that we have included a variety of scores calculated by up to 15 different prediction algorithms^16–25^. For binding algorithms, we included IC50 and percentiles for each peptide candidate predicted against the patient’s HLA allele, and we calculated the median across all algorithms as another feature to be included in the model. Additionally, we have included features representing the physical properties of the peptide, such as the inclusion of cysteines which may affect manufacturability for vaccines.

### Missing data imputation

Missing values were imputed using IterativeImputer (scikit-learn), which iteratively models each feature as a function of all other features until convergence. Features with missing values primarily comprise binding prediction scores and percentiles of wild-type peptides arising from frameshift mutations, with additional sparse missingness in allele expression and RNA VAF features (Supplementary Table 2). For the model described in this manuscript, missing values in NetMHC, SMM, and SMMPMBEC prediction columns were first filled with feature medians prior to imputation. This step was taken because these features had high rates of missingness, and pre-filling with the median reduced the imputation burden on these columns, improving the stability of the IterativeImputer’s internal regression models. The remaining missing values across all other features were then imputed using IterativeImputer. In the implementation integrated into pVACtools v7.0.0, this median pre-filling step was omitted; instead, IterativeImputer was applied directly to all features with missing values across all available samples prior to label-based filtering (see GitHub repository https://github.com/griffithlab/NEAT). Although Review-labeled records were subsequently excluded from model training, their inclusion during imputation improves imputer stability without introducing label leakage, as the imputer relies solely on feature values rather than ITB labels. Across both implementations, 36 features were subject to imputation (Supplementary Table 2), with missing values present in 274 of 1,943 peptide records across 21 patients.

To assess imputation quality, we performed a Missing Completely at Random (MCAR) simulation using complete-case records as ground truth. Fifteen percent of values were randomly masked per feature, and a fresh IterativeImputer instance was fitted exclusively on the masked dataset to prevent leakage of true values. Imputed values were evaluated against masked ground truth using normalized RMSE (NRMSE; RMSE divided by feature standard deviation) and the coefficient of determination (R²) to enable scale-independent comparison across features.

### Selecting ML algorithms for automating ITB decision-making

To evaluate the impact of model architecture and class imbalance strategies on neoantigen candidate classification, we trained both logistic regression and random forest classifiers to predict binary evaluation outcomes (*Accept* vs. *Reject*). Given the class imbalance in our dataset (approximately 25% Accept, 75% Reject), we tested two strategies to mitigate bias toward the majority class: class weighting and random down-sampling.

For logistic regression, models were implemented as scikit-learn pipelines consisting of feature standardization (using StandardScaler) followed by logistic regression. Two separate pipelines were constructed: one using class weighting (class_weight=’balanced’) and another employing random down-sampling using RandomUnderSampler from the imbalanced-learn library, which balances each training fold by reducing the majority class to match the size of the minority class.

Hyperparameter tuning was performed via 5-fold stratified cross-validation using GridSearchCV, optimizing regularization strength (C, ranging from 0.001 to 100), penalty type (L1/Lasso vs. L2/Ridge), and solver (liblinear). Model selection was based on area under the Receiver Operating Characteristic curve (AUROC).

The random forest models were also trained with the same class-balancing strategies. The first used class weighting via scikit-learn’s RandomForestClassifier with class_weight=’balanced_subsample’, which adjusts class weights inversely proportional to class frequencies within each bootstrap sample during tree construction, maintaining the original sample size while penalizing minority class misclassifications. The second used random down-sampling via BalancedRandomForestClassifier from the imbalanced-learn library, which randomly down-samples the majority class in each bootstrap sample before tree construction, creating balanced training subsets for each tree. Both models underwent identical hyperparameter optimization using GridSearchCV with 3-fold cross-validation, searching max_features (mtry) from 1 to 72 and n_estimators (ntree) from 1 to 5000 in steps of 50. Model selection was based on AUROC. The best hyperparameters from grid search were used to train final models with out-of-bag (OOB) scoring enabled.

Performance for these models were evaluated using sensitivity, specificity, F1 scores, AUROC and area under the precision recall curve (AUPR). We selected a final model based on the overall performance of these metrics.

### Making predictions on new cases

The evaluation process during an ITB meeting involves more than a simple binary decision of accepting or rejecting a neoantigen candidate. While many candidates can be readily classified, such as those with high DNA and RNA VAF, strong expression, favorable MHC class I and II binding affinities, and the absence of cysteine residues, each patient typically presents a subset of candidates that require closer scrutiny and are labeled as ‘Review’. These ambiguous cases are manually assessed using pVACview and the Integrative Genomics Viewer (IGV) for reasons that may include: (1) multi-nucleotide mutations or proximal variants in which phasing must be considered to correctly determine protein effect; (2) allele expression for insertion/deletion mutations that may be underestimated due to alignment challenges; (3) variants in transcript isoforms that are poorly expressed or based on low-quality annotations; (4) candidates with poor median binding prediction scores but strong predictions from a subset of individual algorithms; and (5) peptides requiring inspection of anchor residue positions, which influence MHC binding stability.

To streamline the review process, the ML model developed in this study was designed to distinguish straightforward Accept and Reject cases from those genuinely requiring expert attention, thereby reducing the volume of candidates that must be manually assessed. The model outputs a probability score for each candidate, which is used to assign it to one of three categories: Accept, Reject, or Review. Classification thresholds were calibrated using 4 patients from the prospective test set cohort who were not used during model training or development testing. Thresholds were chosen to maximize classification accuracy while minimizing the false negative rate. A candidate is classified as Accept if its predicted probability score exceeds 0.55, Review if the probability falls between 0.3 and 0.55, and Reject if below 0.3. Using these thresholds, we applied the model to all 12 patients in the prospective test set.

### Integration of the model into pVACtools suite

To ensure that this model could be integrated smoothly into our personalized cancer vaccine prioritization workflow as a component of ImmunoNX^15^ the best-performance model has been integrated into the pVACtools suite v7.0.0 as NEAT v1.0.0. The model was implemented to support both integrated execution within the pVACseq pipeline and standalone post-processing. In the integrated mode, model predictions are automatically generated during a pVACseq run when the ‘--run-ml-predictions’ flag is specified. For users operating on existing pVACseq results, a standalone command-line utility, ‘pvacseq add_ml_predictions’, is provided to apply the model independently. This utility accepts the required input files, and outputs the prediction results in the same format as the integrated mode. We allow for customizable classification thresholds for Accept and Reject decisions.

The resulting file is pVACview-compatible, which is a comprehensive interactive visualization tool developed as part of the pVACtools suite for neoantigen analysis and selection during the ITB meeting^12^. Upon loading the predictions into pVACview, ML-based evaluations are automatically populated in the interface, with the predicted probability scores displayed alongside each candidate. These predictions can be reviewed and overridden by users based on additional contextual information available within pVACview, such as binding affinity plots and anchor residue analyses. This integration allows the model to serve as a transparent and interpretable decision-support tool while retaining user flexibility in final candidate selection.

### ITB meeting observation evaluation

In addition to evaluating model predictions against final decisions made during ITB meetings, we sought to assess the model’s impact within a real-world clinical workflow.

Specifically, we observed ITB member responses to the automated model-informed neoantigen decision-making in newly enrolled clinical trial cases. Once raw sequencing data from a patient became available, each sample was processed through the standard ImmunoNX pipeline. We used features generated from the pipeline to make predictions on each candidate and presented them during the corresponding ITB meeting.

This evaluation followed a two-stage design. In the first stage, model predictions were withheld from the board for 4 patient cases in the prospective test set, allowing observation of the standard ITB review process. During this phase, we recorded the time spent reviewing each neoantigen candidate in the absence of model guidance. In the second stage, model predictions were made visible for 3 additional patient cases. During these meetings, we documented both the time spent on candidate review and board member commentary regarding the model’s recommendations. To compare the distribution of ITB review times between the pre- and post-model phases, we applied two complementary statistical tests: a Mann–Whitney U test to assess differences in central tendency, and a two-sample Kolmogorov–Smirnov (KS) test to detect broader differences in distributional shape, spread, or location. Both tests are non-parametric and do not require assumptions of normality or outlier exclusion. Review time distributions were additionally visualized using empirical cumulative distribution functions (ECDFs). The number of patient cases included in this analysis was limited by the rate of enrollment in the ongoing clinical trial during the model’s development period.

## Results

### Cohort assembly

The final dataset comprised 1,943 expert-labeled neoantigen candidates from 33 patients representing 8 cancer types across three independent clinical trials (Table 1.1, Table 1.2). These data were collected over a three-year period and reviewed across 20 ITB meetings, resulting in a uniquely comprehensive resource for training and evaluating a data-driven neoantigen prioritization model. Given that each patient case requires personalized genomic sequencing, HLA typing, and multidisciplinary expert review, curated clinical-grade neoantigen datasets of this scale, diversity, and longitudinal collection are exceptionally rare.

### Selecting ML algorithms for automating ITB decision-making

We compared the performance of two model types, logistic regression and random forest, for predicting neoantigen decision outcomes. Given the class imbalance in the dataset (approximately 25% Accept and 75% Reject), each model was trained using two strategies to address imbalance: class weighting and down-sampling (Table 2). For logistic regression, hyperparameter tuning selected L1 (Lasso) regularization. Under both imbalance strategies, the class-weighted logistic regression model outperformed the down-sampled version across all evaluation metrics. However, overall performance remained modest, with relatively low F1 scores (0.719 and 0.697) and AUPR values (0.738 and 0.685), indicating limited effectiveness in identifying high-priority candidates.

**Table 2.** Performance comparison of models with different class imbalance strategies. Performance metrics for logistic regression and random forest classifiers using either class weighting or down-sampling. All metrics except random forest out-of-bag (OOB) AUROC were evaluated on the development test set. The random forest model with down-sampling achieved the highest F1 score (0.828), highest accuracy and excellent AUROC (0.955) and AUPR (0.899) scores, reflecting strong overall performance in both sensitivity and specificity.

| Model family | Class imbalance strategy | OOB AUROC | Accuracy | Sensitivity | Specificity | F1 | AUROC | AUPR |
| --- | --- | --- | --- | --- | --- | --- | --- | --- |
| Logistic regression | Class weighting | — | 0.816 | 0.859 | 0.8 | 0.719 | 0.894 | 0.738 |
| Logistic regression | Down-sampling | — | 0.803 | 0.824 | 0.796 | 0.697 | 0.868 | 0.685 |
| Random Forest | Class weighting | 0.911 | 0.881 | 0.612 | 0.982 | 0.738 | 0.961 | 0.908 |
| Random Forest | Down-sampling | 0.980 | 0.903 | 0.847 | 0.924 | 0.828 | 0.955 | 0.899 |

In contrast, random forest models consistently outperformed logistic regression across nearly all metrics. Both random forest models were first evaluated using out-of-bag (OOB) AUROC as an internal estimate of generalization performance during training, where the down-sampled model scored notably higher (0.980) than the class-weighted model (0.911). This advantage was confirmed on the held-out development test set, where the down-sampled random forest achieved the best overall performance with higher accuracy (0.903), sensitivity (0.847), and F1 score (0.828). While the class-weighted random forest achieved slightly higher specificity (0.982 vs. 0.924) and AUPR (0.908 vs. 0.899), this came at a substantial cost to sensitivity (0.612 vs. 0.847), reflecting a bias toward the majority Reject class. The improved sensitivity of the down-sampled model is particularly important for prioritizing rare positive (Accept) cases.

Based on this comprehensive evaluation, we selected the random forest model with down-sampling as the final model for downstream testing and integration into the pVACtools pipeline.

### Imputation Quality Assessment

Of the 1,324 peptide records across 21 patients, 274 (20.7%) contained at least one missing value across 36 of 72 features used for model training. Missing values in wild-type (WT) MHC binding and presentation scores arise primarily from frameshift mutations, which do not have a corresponding WT peptide, while missing mutant (MT) scores reflect cases where the given prediction algorithm does not support the patient’s HLA allele. Only 6 features exceeded 10% missingness. To validate imputation quality, we performed an MCAR simulation in which 15% of values were randomly masked from complete-case records and re-imputed using a freshly trained IterativeImputer instance. Imputation performance was strong across the majority of features, with 22 of 36 achieving NRMSE < 0.5 and R² mostly above 0.75 (Supplementary Table 2), particularly for MHC percentile and IC50 scores which had the highest missingness rates. A small subset of features showed poor imputation quality, including RNA VAF, Allele Expression, Corresponding Fold Change, and peptide position (Pos; NRMSE > 1.0 or negative R²). As imputation serves as a prerequisite for model training rather than an end in itself, and Random forest models are inherently robust to moderate imputation noise through ensemble averaging, the downstream validity of the approach is best assessed through held-out predictive performance on the prospective test set.

### Performance of the chosen final model

To evaluate the performance of the selected model random forest with down-sampling, we assessed its ability to classify neoantigen candidates using two complementary approaches: out-of-bag (OOB) evaluation during training and performance on the held-out development test set. During training, the model achieved an OOB AUROC of 0.980, providing an internal estimate of generalization performance on unseen data without requiring a separate validation set. On the held-out development test set, the model achieved strong overall discrimination, with an AUROC of 0.955 and AUPRC of 0.899 (Figure 3B), consistent with the high OOB estimate and confirming that the model generalizes well beyond the training data. The confusion matrix further supports the model’s effectiveness, showing 208 true negatives and 72 true positives, with only 17 false positives and 13 false negatives (Figure 3C), resulting in a 90.3% accuracy, a sensitivity of 0.847, and a specificity of 0.924, highlighting the model’s ability to identify immunogenic neoantigens while minimizing false classifications.

**Figure 3.**
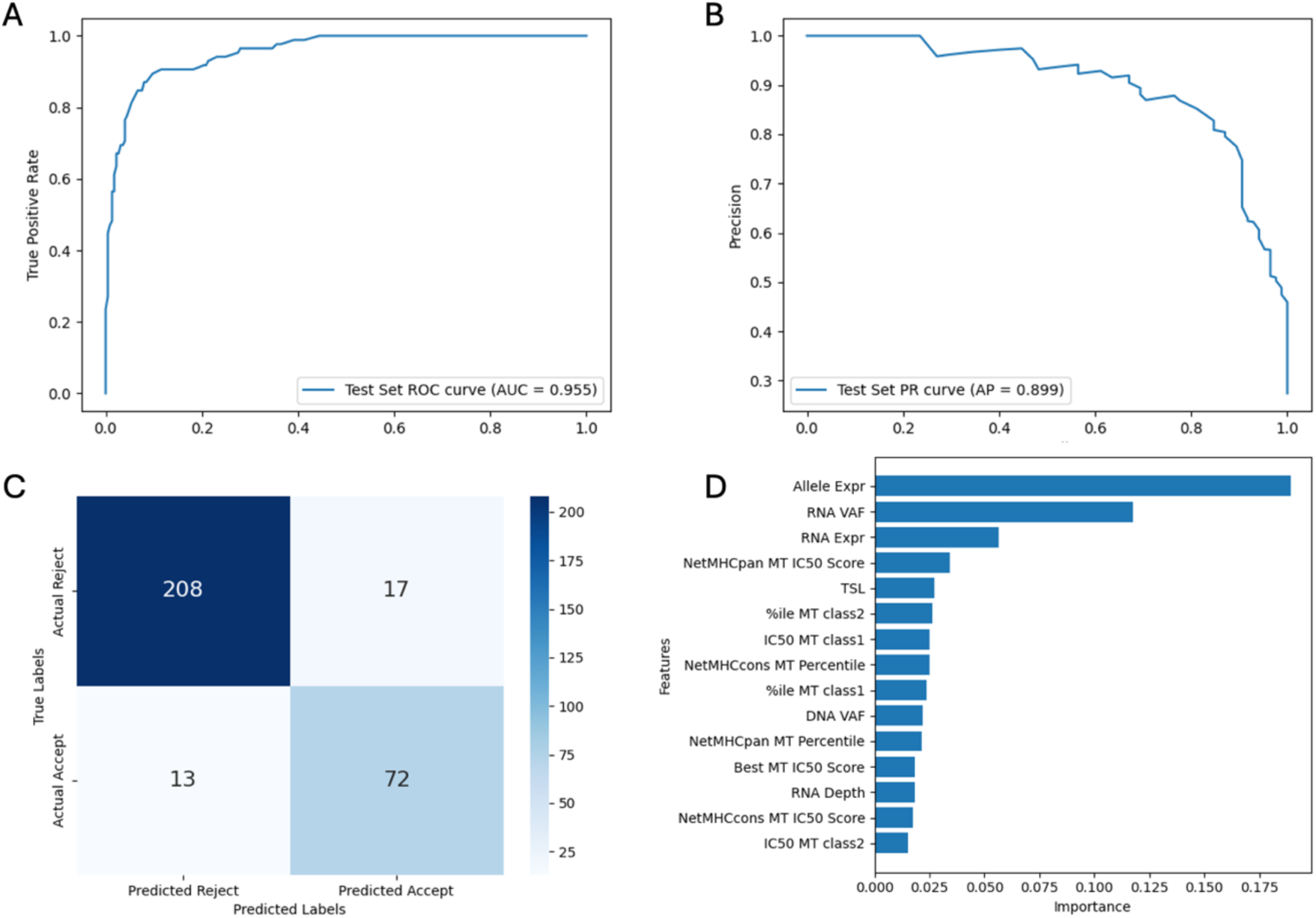
Performance evaluation of the final model (random forest with down-sampling) on the development test set. (A) Receiver Operating Characteristic (ROC) curve of the model on the development test set, demonstrating strong discriminatory ability with an Area Under the ROC (AUROC) of 0.96. (B) Precision-Recall (PR) curve of the same model, with an Area Under the PR Curve (AUPRC) of 0.90, indicating robust performance in the presence of class imbalance. (C) Confusion matrix showing classification outcomes on the development test set. The model correctly classified 208 rejected and 72 accepted neoantigen candidates, with 17 false positives and 13 false negatives. (D) Top 15 most important features (of 72 total features) ranked by Mean Decrease in Impurity (MDI). Features related to RNA expression, allele-specific expression, and predicted MHC binding affinity were among the most influential in determining model predictions.

Feature importance analysis using Mean Decrease in Impurity (MDI) revealed that allele-specific expression, RNA VAF, and RNA expression were the most influential features driving model predictions (Figure 3D). Several features related to MHC binding affinity, including the median IC50 and percentile scores of the mutant peptide predicted across all prediction algorithms and a few individual algorithm predictions, also contributed meaningfully. TSL was ranked as the fifth most important feature, and is often considered during the evaluation process as an indicator for how well-supported the chosen transcript is in Ensembl. Notably, a TSL value greater than 1 is typically flagged by the ITB as “Review” for closer inspection. Tumor DNA VAF was also identified as an informative feature, frequently used to distinguish between clonal and subclonal mutations. RNA depth, in combination with RNA VAF and expression, provides supporting evidence for the abundance of the mutant transcript. Together, these top-ranking features reflect both quantitative and qualitative considerations used during ITB review, underscoring the model’s ability to replicate expert decision-making in neoantigen prioritization.

### Application to the prospective test set

To provide a third and fully independent layer of evaluation beyond OOB estimation and the development test set, we assessed the final model on a prospective test set comprising 12 patients (Table 1.2) whose data were not available at the time of model training or threshold calibration. Using a binary classification framework (Accept vs. Reject), the model achieved an AUROC of 0.948 and an AUPR of 0.794 (Supplementary Figure 1). We then applied predefined probability thresholds (see Methods) to classify all 706 neoantigen candidates into three categories: Accept, Reject, and Review. The resulting performance is shown in the confusion matrix (Figure 4A), with an overall accuracy of 66%. This moderate accuracy is primarily driven by two factors. First, a substantial number of Reject candidates (n = 119) were classified as Review. This reflects the intentionally conservative thresholding strategy, designed to minimize the risk of excluding potentially high-value candidates by shifting uncertain predictions into the Review category. Second, 56 candidates were predicted as Accept but labeled as Reject. This discrepancy likely reflects practical constraints in ITB decision-making, where only a limited number of candidates (typically ∼20) can be selected for vaccine inclusion in patients with high neoantigen burden. Importantly, no true Accept candidates were misclassified as Reject, corresponding to a false negative rate of zero for high-priority neoantigens. Class-specific evaluation using a one-versus-rest approach further demonstrated strong sensitivity, specificity, and F1 scores for the Accept category (Figure 4B). This performance is particularly critical for vaccine design, where maximizing inclusion of truly actionable neoantigens is essential given the limited rate of functional immunogenicity. To confirm that these results were not inflated by inclusion of the 4 patients used for threshold calibration, we repeated this evaluation on the remaining 8 patients (n = 536 peptides); the model maintained strong performance (Supplementary Figure 2).

**Figure 4.**
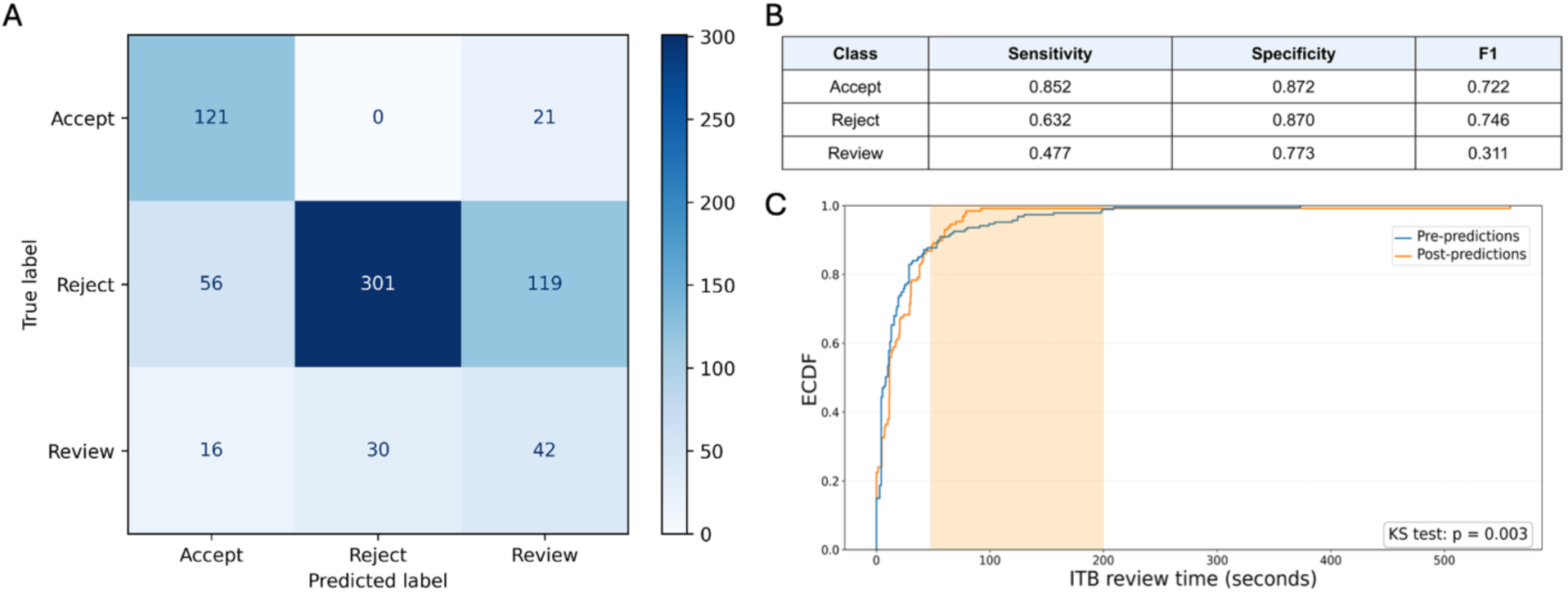
Evaluation of model performance on the prospective test set demonstrating the real-world review setting. (A) Confusion matrix showing the classification results of the final model (random forest with down-sampling) applied to the prospective test set. The model categorized neoantigen candidates into three groups: *Accept*, *Reject* and *Review*. (B) Classification performance of the model evaluated with sensitivity, specificity and F1 using a one-versus-rest strategy. (C) Empirical cumulative distribution function (ECDF) of ITB review time per neoantigen candidate for cases reviewed prior to (Pre-predictions, blue, n=187) and following (Post-predictions, orange, n=129) introduction of model predictions. The shaded region (∼50–200 seconds) highlights the range in which the greatest shift in review time distribution was observed, with a higher proportion of candidates reviewed within this window in the Post-predictions group. A two-sample Kolmogorov-Smirnov test indicated a statistically significant difference between the two distributions (p = 0.003). Extreme outliers (review times >200 seconds) reflecting complex cases requiring extended discussion are included in the statistical test but fall outside the primary region of interest.

During this analysis, we manually inspected a number of candidates that were classified as *Reject* but were in fact *Review* in IGV. During this process, we identified 4 cases in which the mutation expression level was marked as 0 due to a discrepancy of the mutation position called between the DNA aligner and RNA aligner. This issue has been fixed since it has been identified and incorporated into the ImmunoNX pipeline version v1.30 and above (see GitHub Issue #176 [https://github.com/wustl-oncology/analysis-wdls/issues/176]).

Among the 12 patients in the prospective test set, we applied the model to 7 cases to evaluate its practical impact during ITB meetings. Review time per neoantigen candidate was recorded for 4 patients prior to revealing model predictions and for 3 patients after model predictions were made available to board members (n = 187 and n = 129 candidates, respectively; Figure 4C). In both phases, several extreme outliers were observed (review times >200 seconds), reflecting complex cases that required extensive discussion. For example, one case involved a frameshift mutation generating multiple potential neoantigen peptides, while another prompted detailed discussion regarding similarity between the mutated peptide and the human reference proteome. Such cases necessitate manual review irrespective of model implementation.

The ECDF plot (Figure 4C) shows the distributions were significantly different between the Pre- and Post-predictions groups (KS test, p = 0.003). When model predictions were available, a greater proportion of neoantigen candidates were reviewed and resolved within 50–200 seconds compared to the pre-predictions phase. In other words, candidates that previously consumed moderate amounts of discussion time were handled more efficiently (shaded region, Figure 4C). By contrast, candidates resolved very quickly (under 50 seconds) were similarly distributed between groups, suggesting that the model’s benefit was specific to moderately complex candidates rather than those already straightforward enough to decide at a glance. While the overall median review time did not decrease and the Mann-Whitney U test was not significant (p = 0.124), likely reflecting fixed meeting durations and the adjustment period required for board members to incorporate model output into their workflow, the shift in distributional shape indicates that model predictions reduced the time spent on moderately complex candidates that previously required extended discussion. These findings suggest that model-guided decision support streamlines review of routine candidates while retaining thorough expert evaluation for complex cases. Beyond time savings, model-guided pre-classification holds promise for reducing inter-reviewer variability by anchoring discussion around standardized probability scores, directing deliberation toward genuinely complex cases, and improving the overall consistency and scalability of the neoantigen review process.

### Integration to pVACview

To enable seamless use of the trained ML model in clinical and research workflows, we integrated the model into the pVACtools (v7.0.0) suite via two modes: direct execution within the pVACseq pipeline and standalone post-processing (see Methods). Example usage of the integrated pipeline execution is shown in Figure 5A (left), while the standalone post-processing approach is illustrated on the right.

**Figure 5.**
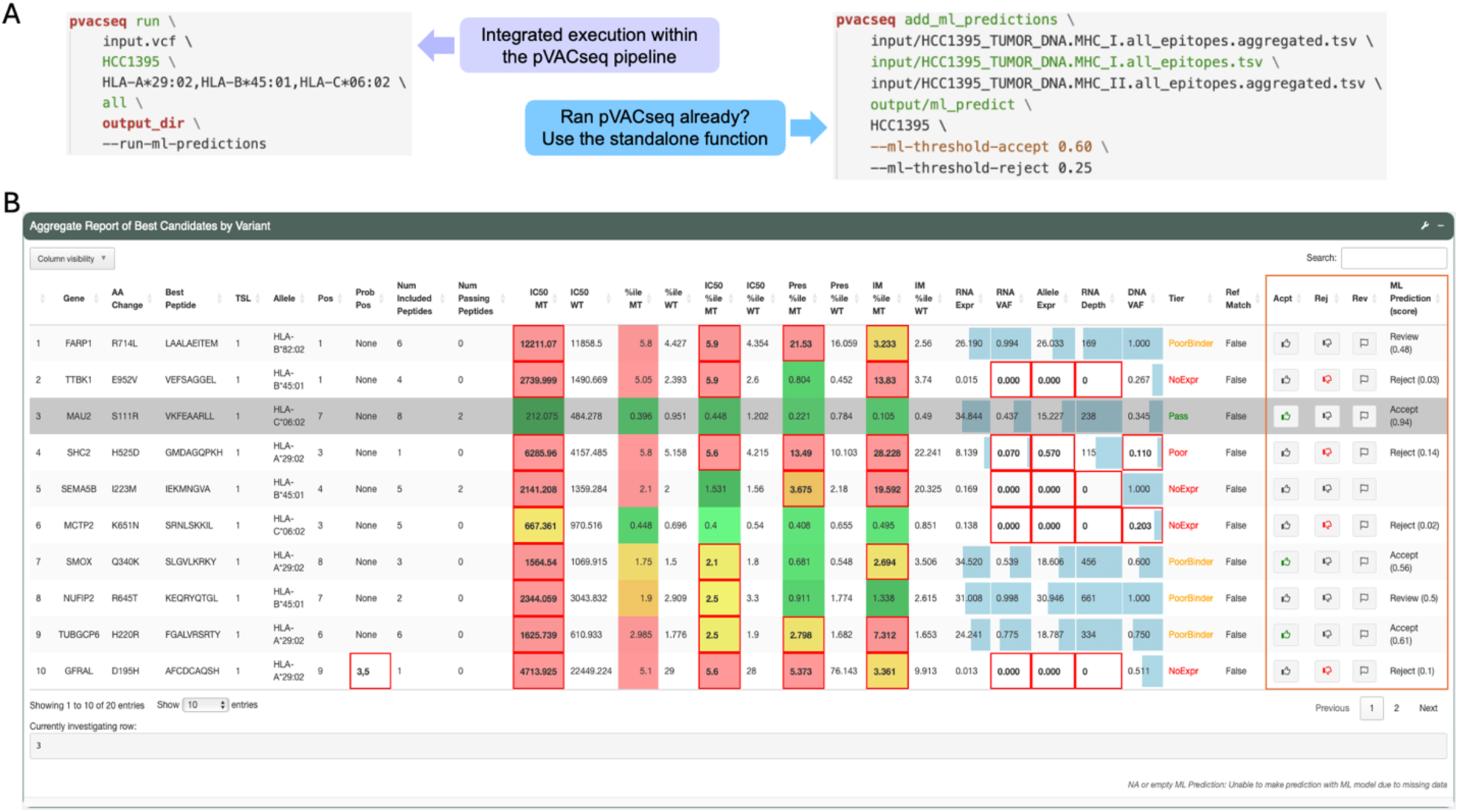
Integration of model into the pVACtools suite. (A) Example command-line usage demonstrating two modes for applying the ML model. Left: integrated execution during a pVACseq run using the ‘--run-ml-predictions’ flag. Right: standalone application to existing pVACseq outputs using the ‘pvacseq add_ml_predictions’ command, with customizable probability thresholds for Accept and Reject classifications. (B) Screenshot of the pVACview interface showing neoantigen candidates with ML model predictions pre-selected. The ML Prediction (score) column displays the model’s categorical recommendation (*Accept*, *Review*, or *Reject*) along with the predicted probability score.

Within pVACview, ML predictions are displayed alongside candidate features, including binding affinity, expression, and variant-level metrics (Figure 5B). Each neoantigen is assigned a categorical recommendation (Accept, Review, or Reject) together with a probability score, allowing users to directly compare model output with manual evaluation. Importantly, these predictions are pre-populated but remain fully editable, supporting a hybrid workflow in which model guidance serves as an initial prioritization step rather than replacing expert review.

## Discussion

As the field of cancer immunotherapy and personalized cancer vaccine development continues to evolve, an increasing number of computational tools and pipelines have emerged to support neoantigen characterization and prioritization. The process of identifying effective neoantigens, those most likely to elicit a robust T-cell response and induce cancer regression, is inherently complex. It requires the integration of multiple biological features, including somatic mutation type, VAF, variant expression, peptide-MHC binding predictions, presentation scores, reference proteome matches, driver gene status, and many others. While individual algorithms have been developed to model discrete components of this workflow, few approaches offer a comprehensive, context-aware method that reflects how experts make decisions in clinical or translational settings.

Within the framework of an ITB, a multidisciplinary team holistically evaluates each neoantigen candidate on a patient-by-patient basis. This expert-driven process, while thorough, is time-intensive and may be subject to human bias and inconsistency. As the number of patients enrolled in PCV trials continues to grow, we sought to address the need for a scalable, systematic, and interpretable approach by developing a ML model to automate and support neoantigen candidate prioritization and decision-making based on historical ITB decisions.

Our model is explicitly trained to replicate ITB decision-making, classifying candidates as *Accept*, *Review*, or *Reject*, based on retrospective data from multiple clinical trial patients. Unlike traditional prioritization methods that rely heavily on static scoring thresholds (e.g. MHC binding affinity <500 nM), our model learns from a set of curated decisions that reflect the nuanced and contextual reasoning of bioinformaticians, clinicians, and immunologists. This includes integrating peptide-level features (binding affinity, agretopicity, reference match) and mutation-level features (allele frequency, genomic context). In doing so, the model captures complex interactions and clinical heuristics that are otherwise difficult to encode manually.

Importantly, the model achieved a false negative rate of zero for high-priority neoantigens, with no true Accept candidates misclassified as Reject. This property is particularly critical in PCV design, where the overall rate of functional immunogenicity is low and the primary objective is to avoid excluding potentially effective candidates. In this context, minimizing false negatives is more important than reducing false positives, as missed candidates represent an irreversible loss of therapeutic opportunity, whereas additional candidates can still be filtered during downstream review. Although the model retains a substantial number of candidates in the Review category, this reflects a deliberate design choice to prioritize sensitivity and preserve uncertain but potentially valuable neoantigens. In clinical scenarios where a sufficient number of high-confidence candidates are available, predicted Accept neoantigens could be advanced directly without further manual review. In such settings, the final selection process could become largely automated, with ITB review focused primarily on candidates assigned to the Review category and other complex cases requiring expert judgment. Overall, this framework supports a flexible, risk-aware prioritization strategy that can automate routine neoantigen selection decisions while preserving expert oversight for uncertain or complex cases, thereby improving both the scalability and reproducibility of PCV design. The fixed probability thresholds used in this study (Accept > 0.55, Review 0.3 – 0.55, Reject < 0.3) represent a pragmatic default calibrated on our clinical cohort; however, we recognize that these cutoffs interact with practical constraints of vaccine design that may vary across patients or trials. In particular, because each PCV typically targets approximately 20 neoantigen candidates, the threshold at which candidates are accepted or flagged for review may reasonably shift depending on a patient’s overall mutation burden; for patients with few high-confidence candidates, relaxing the Accept threshold may be necessary to fill the vaccine, while patients with high neoantigen burden may warrant stricter criteria. This practical reality also influences how model performance is interpreted in the prospective test set, where some candidates labeled Reject by the ITB may reflect capacity constraints rather than true rejection. For this reason, the classification thresholds in pVACtools are fully customizable, allowing clinical teams to adapt the model’s output to the specific context of each patient and trial.

To enhance interpretability and clinical utility, the model was integrated into the pVACtools suite, allowing users to incorporate model predictions directly into their analysis workflow and visualize prediction outcomes within pVACview. This integration enables real-time feedback, supports transparent discussion during ITB meetings, and provides a reproducible framework for prioritizing candidates across clinical trials.

Notably, many of the top-ranked features identified in the feature importance analysis are already readily accessible within the current pVACview interface. In the future, the interface could be further streamlined to emphasize these highly informative features, simplifying manual review while retaining clinically relevant information.

Beyond its predictive value, the development of this model also revealed previously unrecognized issues in the neoantigen analysis pipeline. For example, we identified cases where insertion-induced frame shifts caused discrepancies between DNA- and RNA-based alignment tools, leading to incorrect assessments of transcript expression. This issue previously often required manual inspection in IGV during ITB meetings to rescue high-priority candidates. Through systematic analysis during model training and evaluation, we observed that this misalignment was a recurring source of error. Rather than engineering around it in the model, we corrected this issue directly in the ImmunoNX pipeline, improving the accuracy of future analyses across studies (see GitHub Issue #176 [https://github.com/wustl-oncology/analysis-wdls/issues/176]).

A key limitation of this study is the relatively small number of patients available for model training and evaluation, which reflects the practical realities of PCV development rather than a methodological constraint. Each patient case requires whole-exome sequencing, RNA sequencing, HLA typing, and expert ITB review, a resource-intensive process that means datasets of this scale represent years of coordinated effort across multiple ongoing trials. The dataset of 1,943 expert-labeled neoantigen candidates spanning 33 patients and 8 cancer types constitutes, to our knowledge, one of the largest curated collections derived from real clinical PCV workflows. Nonetheless, the diversity of cancer types represented across three independent clinical trials, combined with strong model performance on a fully held-out prospective test set, supports the generalizability of our approach. At the same time, the model is best understood as a living tool that will continue to improve as additional clinical data become available. As additional patients are enrolled and more ITB decisions are recorded, periodic model retraining is expected to improve classification performance, particularly for underrepresented cancer types and complex cases that currently fall into the Review category. Over time, this iterative learning process may enable increasingly accurate and standardized neoantigen prioritization, further reducing the need for manual review while preserving expert oversight for the most challenging cases.

Taken together, our results demonstrate the feasibility and utility of applying machine learning to model expert-guided neoantigen selection in a clinically grounded and scalable manner. By incorporating multi-level data and modeling real-world decision-making, we bridge a critical gap between computational prediction and translational application in PCV development.

While our model prioritizes candidates likely to be selected for inclusion in a vaccine, it does not directly predict whether a given peptide will elicit a functional T-cell response. The most recent release of pVACtools (v7.0.0) incorporates several immunogenicity prediction algorithms; however, these tools are recent additions and have not yet been routinely incorporated into ITB decision-making, though they may be adopted in future workflows. Predicting immunogenicity remains a central challenge in the field. The limited availability of large-scale, high-quality functional validation datasets has constrained the development of robust models for predicting T-cell activation. Although peptide–MHC binding affinity and presentation metrics are often used as proxies for immunogenicity, these features alone frequently demonstrate low positive predictive value in functional assays and do not account for critical patient-specific factors such as immune microenvironment composition, neoantigen clonality, and HLA allelic variation. As functional assay datasets continue to expand and our understanding of T-cell responses, our next objective is to develop an integrative machine learning framework that predicts not only which neoantigens are likely to be selected, but also which are truly immunogenic. Such efforts will further enhance the role of computational modeling in guiding PCV design.

## Data Availability

All relevant data are within the paper and supplementary data. The NEAT (Neoantigen Evaluation & Automated Triage) model, associated code, and manuscript reproducibility notebooks are available at https://github.com/griffithlab/NEAT (v1.0.0).

## Availability

All relevant data are within the paper and supplementary data. The NEAT (Neoantigen Evaluation & Automated Triage) model, associated code, and manuscript reproducibility notebooks are available at https://github.com/griffithlab/NEAT (v1.0.0). Questions and requests may be submitted via the GitHub issue tracker. Documentation on the usage of the model and integration into pVACtools are available at https://pvactools.readthedocs.io/en/7.0.0_docs/. The release number of this model at the time of submission is v1.0.0, coupled with pVACtools v7.0.0.

## Acknowledgments

We are grateful to the patients and their families for the donation of their samples and participation in clinical trials. M.G., O.L.G. and W.E.G were supported by the NIH National Cancer Institute (NCI) under award number P50CA272213. M.G., O.L.G., H.X., W.E.G., and T.A.F. were supported by the NCI under award number U01CA248235. T.A.F and W.E.G were supported by the Siteman Cancer Center (P30CA91842). T.A.F was also supported by the Leukemia SPORE (P50CA171063), Lymphoma Research Foundation, Blood Cancer United, the Howard Steinberg Family Foundation, and the Paula and Rodger Riney Blood Cancer Initiative. M.G., O.L.G. and W.E.G. were supported by the NCI/Leidos Biomedical Research Subcontract 20X012F5P50. S.P.G. was supported by the Foundation for Barnes-Jewish Hospital. K.S. was supported by NIH/NIGMS T32GM139774. JAF was supported by the NCI under K22CA282364 and the Cancer Research Foundation (CRF) Young Investigator Award. W.E.G was supported by Susan G. Komen for the Cure (KG111025), B101/Centene Corporation P19-00559, Alvin J. Siteman Cancer Center Investment Program grant 4035, CA/NCI NIH R01CA240983, MedImmune AstraZeneca 004135-946897 and P50CA196510. This work was also supported in part by the Washington University Institute of Clinical and Translational Sciences from the National Center for Advancing Translational Sciences (NCATS) of NIH under award number UL1TR002345. Finally, this work was supported by a gift from the Goldberg Family Foundation.

