## Supplementary Tables for "Automating neoantigen selection for personalized cancer vaccine design"

**Supplementary Table 1 List of all features included in the model**

| Category | Features | Description |
|---|---|---|
| Mutation qualities | DNA VAF | Tumor DNA variant allele frequency (VAF) at this position. |
| | RNA VAF | Tumor RNA variant allele frequency (VAF) at this position. |
| | RNA Expr | Gene expression value for the annotated gene containing the variant. |
| | RNA Depth | Tumor RNA depth at this position. |
| | Allele Expr | RNA Expr * RNA VAF |
| | Pos | Mutation position (A list of all amino acid positions in the MT Epitope Seq that are different from the WT Epitope Seq.) |
| | Variant Type | The type of variant. missense for missense mutations, inframe_ins for inframe insertions, inframe_del for inframe deletions, and FS for frameshift variants |
| | Gene of Interest | Whether the mutated gene is a cancer driver gene for the given cancer type |
| MHC Class I Binding Affinity Prediction Algorithm | NetMHCpan WT IC50 Score | IC50 binding score of the wild-type peptide predicted from NetMHCpan |
| | NetMHCpan MT IC50 Score | IC50 binding score of the mutant peptide predicted from NetMHCpan |
| | NetMHCpan WT Percentile | Percentile rank of the wild-type peptide predicted from NetMHCpan |
| | NetMHCpan MT Percentile | Percentile rank of the mutant peptide predicted from NetMHCpan |
| | NetMHC WT IC50 Score | IC50 binding score of the wild-type peptide predicted from NetMHC |
| | NetMHC MT IC50 Score | IC50 binding score of the mutant peptide predicted from NetMHCpan |
| | NetMHC WT Percentile | Percentile rank of the wild-type peptide predicted from NetMHC |
| | NetMHC MT Percentile | Percentile rank of the mutant peptide predicted from NetMHC |
| | NetMHCcons WT IC50 Score | IC50 binding score of the wild-type peptide predicted from NetMHCcons |
| | NetMHCcons MT IC50 Score | IC50 binding score of the mutant peptide predicted from NetMHCcons |

| | | |
|---|---|---|
| | NetMHCcons WT Percentile | Percentile rank of the wild-type peptide predicted from NetMHCcons |
| | NetMHCcons MT Percentile | Percentile rank of the mutant peptide predicted from NetMHCcons |
| | PickPocket WT IC50 Score | IC50 binding score of the wild-type peptide predicted from PickPocket |
| | PickPocket MT IC50 Score | IC50 binding score of the mutant peptide predicted from PickPocket |
| | PickPocket WT Percentile | Percentile rank of the wild-type peptide predicted from PickPocket |
| | PickPocket MT Percentile | Percentile rank of the mutant peptide predicted from PickPocket |
| | SMM WT IC50 Score | IC50 binding score of the wild-type peptide predicted from SMM |
| | SMM MT IC50 Score | IC50 binding score of the mutant peptide predicted from SMM |
| | SMM WT Percentile | Percentile rank of the wild-type peptide predicted from SMM |
| | SMM MT Percentile | Percentile rank of the mutant peptide predicted from SMM |
| | SMMPMBEC WT IC50 Score | IC50 binding score of the wild-type peptide predicted from SMMPMBEC |
| | SMMPMBEC MT IC50 Score | IC50 binding score of the mutant peptide predicted from SMMPMBEC |
| | SMMPMBEC WT Percentile | Percentile rank of the wild-type peptide predicted from SMMBEC |
| | SMMPMBEC MT Percentile | Percentile rank of the mutant peptide predicted from SMMBEC |
| | MHCflurry WT IC50 Score | IC50 binding score of the wild-type peptide predicted from MHCflurry |
| | MHCflurry MT IC50 Score | IC50 binding score of the mutant peptide predicted from MHCflurry |
| | MHCflurry WT Percentile | Percentile rank of the wild-type peptide predicted from MHCflurry |
| | MHCflurry MT Percentile | Percentile rank of the wild-type peptide predicted from MHCflurry |
| | MHCnuggetsI WT IC50 Score | IC50 binding score of the wild-type peptide predicted from MHCnuggetsI |
| | MHCnuggetsI MT IC50 Score | IC50 binding score of the mutant peptide predicted from MHCnuggetsI |
| | MHCnuggetsI WT Percentile | Percentile rank of the wild-type peptide predicted from MHCnuggetsI |

| | | |
|---|---|---|
| | MHCnuggetsI MT Percentile | Percentile rank of the mutant peptide predicted from MHCnuggetsI |
| MHC Class I Elution Prediction Algorithm | NetMHCpanEL WT IC50 Score | IC50 binding score of the wild-type peptide predicted from NetMHCpanEL |
| | NetMHCpanEL MT IC50 Score | IC50 binding score of the mutant peptide predicted from NetMHCpanEL |
| | NetMHCpanEL WT Percentile | Percentile rank of the wild-type peptide predicted from NetMHCpanEL |
| | NetMHCpanEL MT Percentile | Percentile rank of the mutant peptide predicted from NetMHCpanEL |
| | MHCflurryEL Presentation WT Score | Presentation score of the wild-type peptide predicted from MHCflurryEL |
| | MHCflurryEL Presentation MT Score | Presentation score of the mutant peptide predicted from MHCflurryEL |
| | MHCflurryEL Presentation WT Percentile | Percentile rank of the wild-type peptide predicted from MHCflurryEL |
| | MHCflurryEL Presentation MT Percentile | Percentile rank of the mutant peptide predicted from MHCflurryEL |
| | MHCflurryEL Processing WT Score | Processing score of the wild-type peptide predicted from MHCflurryEL |
| | MHCflurryEL Processing MT Score | Processing score of the mutant peptide predicted from MHCflurryEL |
| MHC Class I summary scores | IC50 WT class1 | Median IC50 binding affinity of the corresponding wild-type epitope across all Class I prediction algorithms used |
| | IC50 MT class1 | Median IC50 binding affinity of the corresponding mutant epitope across all Class I prediction algorithms used |
| | %ile WT class1 | Median binding affinity percentile rank of the best-binding wild-type epitope across all Class I prediction algorithms used (those that provide percentile output) |
| | %ile MT class1 | Median binding affinity percentile rank of the best-binding mutant epitope across all Class I prediction algorithms used (those that provide percentile output) |
| | Best MT IC50 Score | Lowest IC50 binding affinity of all Class I prediction algorithms used |
| | Corresponding WT IC50 Score | IC50 binding affinity of the wild-type epitope that corresponds to the best MT IC50 Score. |
| | Best MT Percentile | Lowest percentile rank of all Class I prediction algorithms used |

| | | |
|---|---|---|
| | Corresponding WT Percentile | Binding affinity percentile rank of the wild-type epitope that corresponds to the best MT Percentile. |
| | Median Fold Change | IC50 WT class1 / IC50 MT class1 |
| | Corresponding Fold Change | Corresponding WT IC50 Score / Best MT IC50 Score |
| MHC Class II summary scores | IC50 WT class2 | Median IC50 binding affinity of the corresponding wild-type epitope across all Class II prediction algorithms used |
| | IC50 MT class2 | Median IC50 binding affinity of the corresponding mutant epitope across all Class II prediction algorithms used |
| | %ile WT class2 | Median binding affinity percentile rank of the best-binding wild-type epitope across all Class II prediction algorithms used (those that provide percentile output) |
| | %ile MT class2 | Median binding affinity percentile rank of the best-binding mutant epitope across all Class II prediction algorithms used (those that provide percentile output) |
| Other | Biotype | The biotype of the affected transcript |
| | Ref Match | Was there a match of the mutated peptide sequence to the reference proteome? |
| | TSL | The transcript support level (TSL) of the affected transcript. Not Supported if the VCF entry doesn’t contain TSL information. |
| | Peptide Length | The peptide length of the epitope |
| | cysteine_count | Number of Cysteines in the amino acid sequence. Problematic because they can form disulfide bonds across distant parts of the peptide |
| | Prob match | Whether the mutated amino acid is a Cystein. |
| | Prob Pos | A list of positions in the Best Peptide that are problematic. |
| | Num Passing Peptides | The number of included peptides for this mutation that are well-binding. |

**Supplementary Table 2 Missing Completely at Random (MCAR) simulation results**

| Feature | N missing | N rows | % missing | NRMSE | R2 |
|---|---|---|---|---|---|
| SMM WT Percentile | 255 | 1324 | 0.193 | 0.411 | 0.841 |
| SMMPMBEC WT Percentile | 255 | 1324 | 0.193 | 0.251 | 0.922 |
| SMM MT Percentile | 219 | 1324 | 0.165 | 0.358 | 0.857 |
| SMMPMBEC MT Percentile | 219 | 1324 | 0.165 | 0.334 | 0.765 |
| NetMHC WT Percentile | 174 | 1324 | 0.131 | 0.279 | 0.896 |
| NetMHC MT Percentile | 135 | 1324 | 0.102 | 0.488 | 0.797 |
| Corresponding Fold Change | 43 | 1324 | 0.032 | 0.539 | -0.448 |
| MHCflurryEL Processing WT Score | 43 | 1324 | 0.032 | 0.498 | 0.702 |
| PickPocket WT IC50 Score | 43 | 1324 | 0.032 | 0.522 | 0.756 |
| SMMPMBEC WT IC50 Score | 43 | 1324 | 0.032 | 0.125 | 0.191 |
| MHCnuggetsI WT IC50 Score | 43 | 1324 | 0.032 | 0.585 | 0.663 |
| NetMHCpan WT Percentile | 43 | 1324 | 0.032 | 0.198 | 0.943 |
| NetMHCpanEL WT Percentile | 43 | 1324 | 0.032 | 0.693 | 0.668 |
| Corresponding WT IC50 Score | 43 | 1324 | 0.032 | 0.799 | 0.576 |
| SMM WT IC50 Score | 43 | 1324 | 0.032 | 0.981 | 0.772 |
| Median Fold Change | 43 | 1324 | 0.032 | 1.046 | 0.249 |
| MHCnuggetsI WT Percentile | 43 | 1324 | 0.032 | 0.662 | 0.613 |
| NetMHCpanEL WT IC50 Score | 43 | 1324 | 0.032 | 0.41 | 0.845 |
| MHCflurryEL Presentation WT Percentile | 43 | 1324 | 0.032 | 0.384 | 0.887 |
| IC50 WT class1 | 43 | 1324 | 0.032 | 0.301 | 0.94 |
| %ile WT class1 | 43 | 1324 | 0.032 | 0.233 | 0.964 |
| NetMHCcons WT Percentile | 43 | 1324 | 0.032 | 0.253 | 0.94 |
| NetMHCcons WT IC50 Score | 43 | 1324 | 0.032 | 0.292 | 0.898 |
| MHCflurry WT IC50 Score | 43 | 1324 | 0.032 | 0.366 | 0.847 |
| MHCflurryEL Presentation WT Score | 43 | 1324 | 0.032 | 0.297 | 0.911 |
| NetMHCpan WT IC50 Score | 43 | 1324 | 0.032 | 0.321 | 0.919 |
| NetMHC WT IC50 Score | 43 | 1324 | 0.032 | 0.325 | 0.898 |

| | | | | | |
|---|---|---|---|---|---|
| MHCflurry WT Percentile | 43 | 1324 | 0.032 | 0.335 | 0.824 |
| PickPocket WT Percentile | 43 | 1324 | 0.032 | 0.365 | 0.77 |
| Corresponding WT Percentile | 43 | 1324 | 0.032 | 1.327 | 0.254 |
| IC50 WT class2 | 34 | 1324 | 0.026 | 0.808 | 0.504 |
| %ile WT class2 | 34 | 1324 | 0.026 | 0.339 | 0.87 |
| Allele Expr | 14 | 1324 | 0.011 | 0.617 | -5.182 |
| RNA VAF | 14 | 1324 | 0.011 | 1.132 | -0.491 |
| RNA Depth | 14 | 1324 | 0.011 | 1.197 | 0.376 |
| Pos | 1 | 1324 | 0.001 | 1.081 | -0.003 |
