## Supplementary Figures for "Automating neoantigen selection for personalized cancer vaccine design"

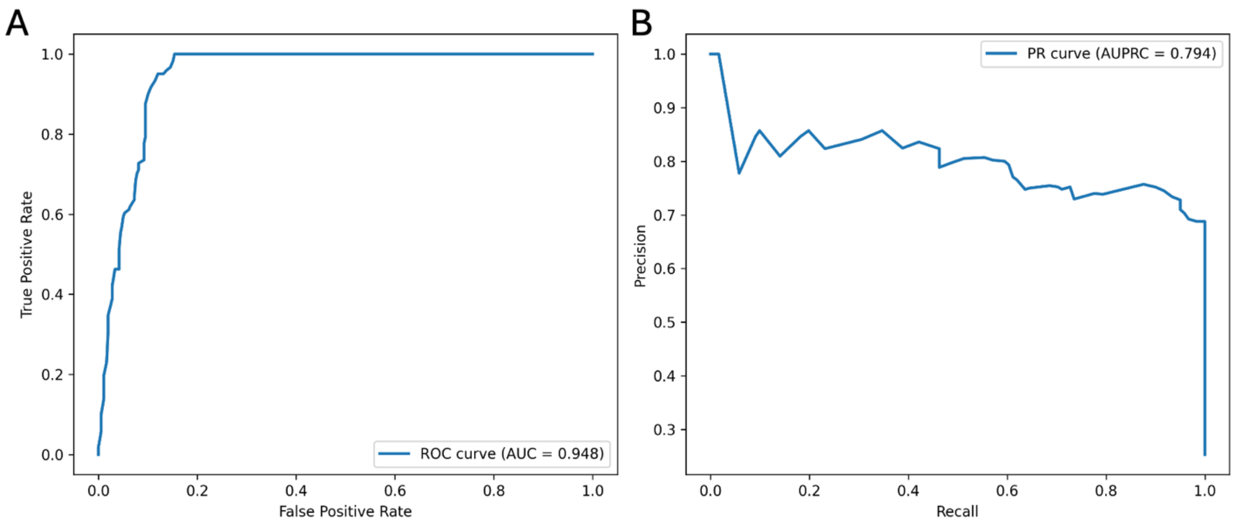


**Supplementary Figure 1. Binary classification performance of the final model on the prospective test set.**

(A) Receiver Operating Characteristic (ROC) curve evaluating the ability of the final model (Random Forest with down-sampling) to discriminate between Accept and Reject neoantigen candidates in the prospective test set (n = 706 peptides, 12 patients). The area under the ROC curve (AUROC = 0.948) indicates strong discriminatory performance.

(B) Precision-Recall (PR) curve for the same model and dataset. The area under the PR curve (AUPRC = 0.794) demonstrates robust performance in the context of class imbalance.

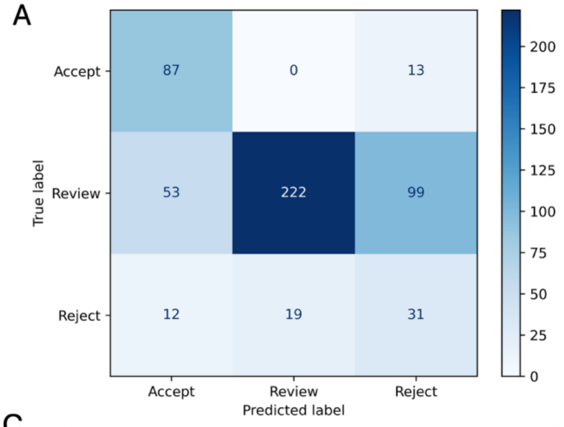


B

| Class | Sensitivity | Specificity | F1 |
|---|---|---|---|
| Accept | 0.870 | 0.851 | 0.690 |
| Reject | 0.594 | 0.883 | 0.722 |
| Review | 0.500 | 0.764 | 0.302 |

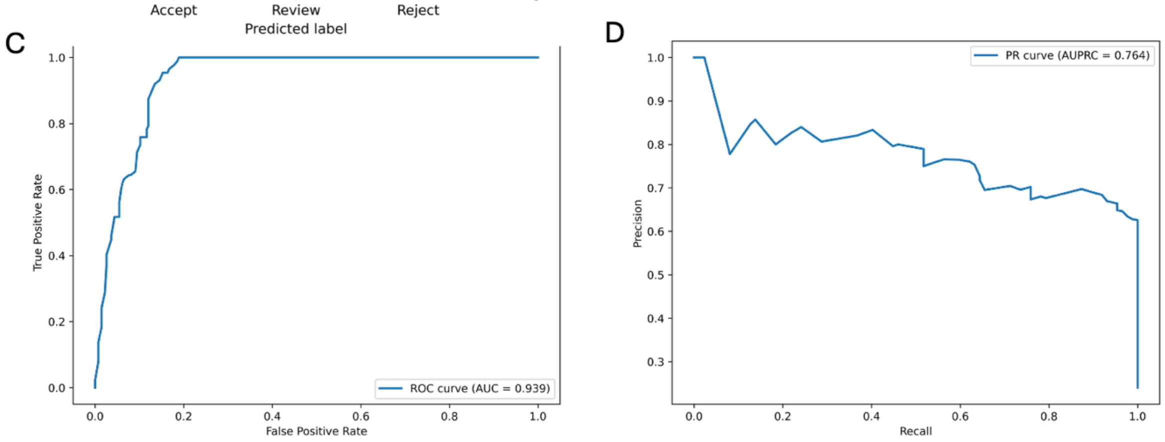


**Supplementary Figure 2. Model performance on the prospective test set excluding threshold calibration patients (n = 536 peptides, 8 patients).**

(A) Confusion matrix showing three-class classification results (Accept, Review, Reject) for the final model applied to the prospective test set after excluding the 4 patients used for probability threshold calibration.

(B) Class-specific performance metrics evaluated using a one-versus-rest strategy, reporting sensitivity, specificity, and F1 score for each category. The Accept class achieved the highest sensitivity (0.870) and specificity (0.851), reflecting the model's prioritization of high-value neoantigen candidates.

(C) ROC curve for binary (Accept vs. Reject) classification on the calibration-excluded prospective test set, with an AUROC of 0.939, demonstrating strong discriminatory performance comparable to the full prospective test set evaluation (AUROC = 0.948; Supplementary Figure 1).

(D) Precision-Recall curve for the same binary classification, with an AUPRC of 0.764. Sustained precision above 0.65 across a broad recall range indicates robust identification of Accept candidates in the presence of class imbalance.
